# Genetic Polymorphisms and Post-Stroke Upper Limb Motor Improvement – A Systematic Review and Meta-Analysis

**DOI:** 10.1101/2023.12.06.23299579

**Authors:** Sandeep K. Subramanian, Riley T. Morgan, Carl Rasmusson, Kayla M. Shepherd, Carol L Li

## Abstract

**Background:** Post-stroke upper limb (UL) motor improvement is associated with adaptive neuroplasticity and motor learning. Both intervention-related (including provision of intensive, variable, and task-specific practice) and individual-specific factors (including the presence of genetic polymorphisms) influence improvement. In individuals with stroke, most commonly, polymorphisms are found in Brain Derived Neurotrophic Factor (BDNF), Apolipoprotein (APOE) and catechol-O-methyltransferase (COMT). These involve a replacement of cystine by arginine (APOEε4) or one or two valines by methionine (BDNF: val^66^met, COMT: val^158^met). However, the implications of these polymorphisms on post-stroke UL motor improvement specifically have not yet been elucidated.

**Objective:** Examine the influence of genetic polymorphism on post-stroke UL motor improvement.

**Design:** Systematic Review and Meta-Analysis

**Methods:** We conducted a systematic search of the published literature in English language of using standard methodology. The modified Downs and Black checklist helped assess study quality. We compared change in UL motor impairment and activity scores between individuals with and without the polymorphisms. Meta-analyses helped assess change in motor impairment scores based upon a minimum of two studies per time point. Effect sizes (ES) were quantified based upon the Rehabilitation Treatment Specification System as follows: small (0.08–0.18), medium (0.19–0.40) and large (≥0.41).

**Results:** We retrieved 10 (four good and six fair quality) studies. Compared to those with BDNF val^66^met polymorphism, meta-analyses revealed lower motor impairment scores (large ES) in those without the polymorphism at intervention completion (0.5, 95% CI: 0.11-0.88) and at retention (0.58, 95% CI: 0.06-1.11). Presence of CoMT val^158^met polymorphism had similar results, with higher levels of improvement in impairment (large ES ≥1.5) and activity scores (large ES ranging from 0.5-0.76) in those without the polymorphism. Presence of APOEε4 form did not influence UL motor improvement.

**Conclusion:** BDNF val^66^met and COMT val^158^met polymorphisms negatively influence UL motor improvement in impairment and activity scores.

**Registration:** https://osf.io/wk9cf/

## Introduction

Stroke continues to be a leading cause of adult morbidity in the United States.^1^ One of the most disabling aftereffects of a stroke is the presence of upper limb (UL) hemiparesis. A large proportion of stroke survivors present with UL sensorimotor impairments on the paretic side, reduced independence in performance of daily life activities (ADL) and restricted participation.^2^ Along with spontaneous recovery mechanisms,^3^ motor improvement of the paretic side enabling successful task-performance is attributable to adaptive neuroplasticity and motor learning.^4^

Successful task-performance entails an interaction of the individual, environment, and the task to be performed.^5^ The role of the environment^6,7^ and factors influencing task-practice^8^ have been extensively studied. Recently, there is a renewed focus on the role of individual-specific characteristics such as levels of motivation,^9,10^ mood^10^ and the role of biomarkers.^11^ Bernhardt et al^12^ defined biomarkers as “*indicators of disease state that can be used clinically as a measure reflecting underlying molecular and cellular processes that may be difficult to measure directly in humans and could be used to predict recovery or treatment response.”* Biomarker studies within the realm of neurorehabilitation include those based on biology (e.g., genetics), structural and/or imaging^13^ and neurophysiological markers^14^ of central nervous system excitability and electrical activity.

The role of imaging-based biomarkers of structural and functional corticospinal tract connectivity alone^13^ or in combination with neurophysiological markers (e.g. motor evoked potential amplitude)^14^ has been extensively studied. The role of genetics-based biomarkers is slowly gaining prominence,^15^ with studies focusing on single nucleotide polymorphisms (SNPs).^11^ These SNPs can alter the basic functioning in cellular and molecular processes^16^ and tend to influence functional improvement produced by environmental interaction and in response to rehabilitation interventions.^17^ Genetics-based biomarkers identified as pertinent to stroke recovery include SNPs in brain-derived neurotrophic factor (BDNF), Catechol-o-methyltransferase (COMT) and Apolipoprotein (APOE).^11^

An activity dependant^18^ neurotrophin important for neuroplasticity and protection after injury, BDNF facilitates synaptic transmission and long-term potentiation important for motor learning.^19^ A common SNP that occurs in BDNF is substitution of one or two valines at codon 66 with methionine due to substitution of adenine in place of guanine at nucleotide 196.^20^ The polymorphism reduces activity-dependent BDNF release,^21^ and results in altered neuroplasticity and learning in healthy controls^22^ and after a stroke.^23,24^

The COMT enzyme helps degrade and thus influences the availability of Dopamine in the central nervous system.^25^. Dopamine can influence post-stroke motor learning and improvement.^26,27^ A commonly observed SNP results in a change from valine or methionine at codon 158 (in the membrane form) and codon 108 in the soluble form, which results in a 3-4 fold decrease in COMT activity.^28,29^ The role of COMT polymorphism has primarily been assessed on motor learning in Parkinson’s disease^30,31^ and severe Schizophrenia.^32^ Given that COMT is found in areas essential for motor learning,^33^ such as striatum and motor cortex,^34^ the effects of COMT polymorphism on post-stroke motor improvement need to be addressed.

Although involved in lipid transport between cells, APOE helps modulate neuronal repair and regeneration of nervous tissue. One of the alleles of APOE is the Epsilon-4 form (ε4) with arginine at positions 112 and 158 in place of cystine. Presence of APOE-ε4 can cause reduced hippocampal volume and cortical thickness, cognitive impairments^35^ and lower recovery levels after traumatic brain^36^ and spinal cord^37^ injuries. After a stroke, previous meta-analyses^38,39^ revealed lower improvement after sub-arachnoid hemorrhage in those with the ε4 form, but no association with improvements after ischemic strokes. In both studies,^38,39^ motor improvements were assessed using generic scales such as the modified Rankin Scale (mRS). Improved scores in assessments such as the mRS does not specifically represent UL motor improvement.^40^ As presence of cognitive impairments influence UL motor improvement,^41^ the effects of the APOE-ε4 form on post-stroke UL motor improvements needs to be systematically evaluated.

The role of genetic polymorphisms has previously been reviewed.^11,16,24,39,42,43^ These studies were either narrative reviews^11,16,24,43^ or meta-analyses including global stroke outcomes like National Institutes of Health Stroke Scale and/or mRS.^39,42^ Post-stroke UL motor improvement continues to remain variable and less than optimal in many cases.^44^ Evaluation of whether and to what extent genetic polymorphisms influence the extent of improvement may help explain some of the observed variability. Using a systematic review and meta-analysis, we examined the influence of genetic polymorphisms on UL motor improvement. The question guiding our review was “*In individuals with post-stroke UL hemiparesis, does the presence, compared to the absence of genetic polymorphisms, influence motor improvement?*” Preliminary results have previously appeared as an abstract.^45^

## Methods

This systematic review followed the PRISMA (Preferred Reporting Items for Systematic Reviews and Meta-Analyses) guidelines. The protocol was registered on the Open Science Framework (https://osf.io/wk9cf/).

We searched the literature for studies involving human subjects published in English between the years 2000 and 2023. The last search was conducted in September 2023. Key search terms used included: stroke, cerebrovascular accident, upper limb, arm, rehabilitation, impairment, activities of daily living, recovery, polymorphisms, gene*, neuroplasticity, and motor learning. Databases searched included: PubMed and ISI Web of Science and the Google Scholar repository. We included studies that used clinical assessments of UL motor impairment and/or ADL and provided data for individuals with and without polymorphisms. We excluded studies focusing exclusively on lower limb or on only cognitive outcomes. We also excluded other reviews, although we searched the reference lists of these excluded reviews for pertinent citations. To identify additional relevant articles, we also searched reference lists of each retrieved study.

### Data Abstraction

We grouped the retrieved articles according to the polymorphism examined. We developed and used a data abstraction form to extract data from the selected articles. Data were initially extracted by RTM, CR and KMS. The first author (SKS) then verified that all relevant data were obtained from the selected articles. The extracted data included details about chronicity, distribution of sample based upon those with and without polymorphism, details about the intervention, outcomes used to assess change and the study results.

### Study quality assessment

We assessed the quality of the selected articles using the modified version^46^ of the reliable and valid Downs and Black (D&B) checklist.^47^ The D&B checklist can be used to assess the quality of both randomized and non-randomized study designs. The total scores of this assessment and PEDro scale are highly correlated in studies involving post-stroke participants.^48^ According to available guidelines,^49^ we classified the scores as “excellent” (score 24-28), “good” (score 19-23), “fair” (score 14-18), or “poor” (score ≤ 13). The quality of each study was independently evaluated by RTM, CR and KMS, with discrepancies, if any, resolved by SKS and CLL.

### Risk of Bias

The risk of bias (ROB) was estimated using the Cochrane ROB tool^50^ and ACROBAT-NRSI (A Cochrane Risk Of Bias Assessment Tool: for Non-Randomized Studies of Interventions)^51^ for randomized and non-randomized studies respectively. The Cochrane ROB tools assesses the following domains: sequence generation, allocation, concealment, blinding of participants, personnel and outcome assessors, incomplete outcome data, selective outcome reporting, and other sources of bias. For each domain, we assigned a judgment: Yes - indicating low ROB, No - indicating a high ROB, and Unclear - indicating unclear or unknown ROB where reported details were insufficient to reach a conclusion. The ACROBAT-NRSI tool assesses bias that can arise because of confounding, study participant selection, intervention measurement, departures from intended interventions, missing data, outcome measurement and reported result selection.

### Statistical analyses

Descriptive statistics of the study populations were calculated as percentages of the total sample. When an article reported the effect of a particular polymorphism at both the motor impairment and activity limitation levels, they were considered separately. Meta-analyses (RevMan 5) examined differences in Fugl-Meyer (FM) scores in groups with and without polymorphism. Pooled effects of the polymorphisms were quantified with standardized mean differences.^52^ If at least two studies reported the effects of the polymorphism on change in FM scores, we included them in the meta-analysis.^53,54^ I^2^ scores helped assess heterogeneity.^55^

Given that a variety of interventions were employed in the different studies, we used the random effects models (irrespective of I^2^ values). Effect sizes were categorized as small (0.08 - 0.18), medium (0.19 - 0.40) and large (≥0.41), in accordance with the Rehabilitation Treatment Specification System recommendations.^56^ Sensitivity analysis was carried out to assess the effect of provision of rehabilitation interventions. We conducted an additional analysis excluding any study that did not report details of rehabilitation interventions provided.

## Results

The search and selection results are shown in the Preferred Reporting Items for Systematic Reviews and Meta-Analyses (PRISMA) flow diagram. In total, 319 citations were identified through database and registry searches (Figure 1). After removing duplicates, 187 citations were screened, of which 16 were excluded. We sought 116 reports for retrieval and assessed 31 for full text eligibility, which were experimental studies including outcomes related to rehabilitation. We further excluded 21 studies, as they included lower limb and/or gait outcomes or used generic measures such as the mRS, NIHSS and Barthel Index. Ten articles assessing the effects of genetic polymorphisms on UL motor impairment and ADL performance were included in the qualitative synthesis (Figure 1). The reference lists of these ten articles did not yield any additional citations.

**Figure 1.**
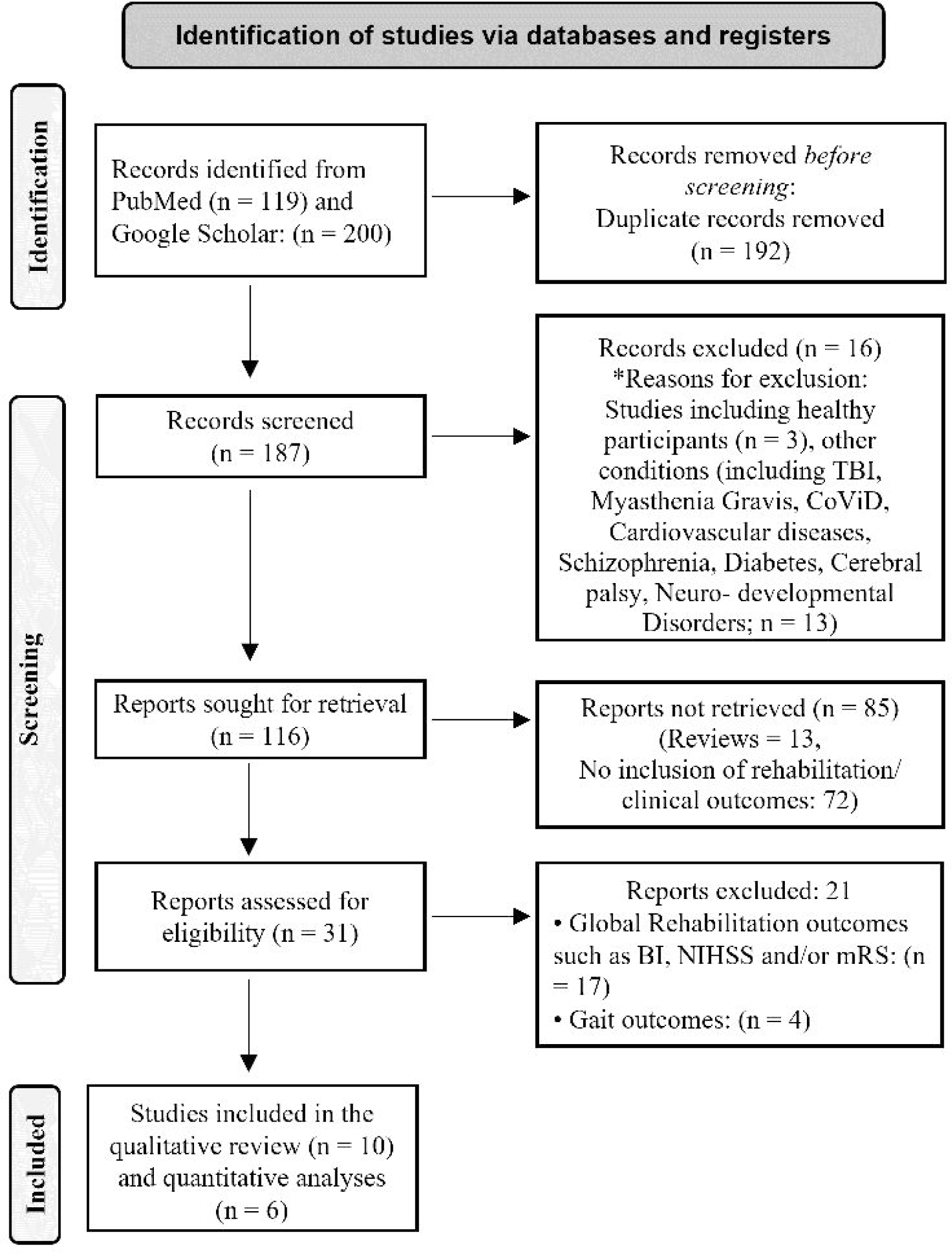
Preferred Reporting Items for Systematic Reviews and Meta-Analyses (PRISMA) flow diagram.

Out of the 10 studies, eight examined the influence of BDNF val^66^met and two addressed the effects of COMT val^158^met polymorphisms. Two of the eight studies assessing the effects of BDNF val^66^met polymorphism additionally examined effects of APOE ε4 polymorphism. Six^23, 57–61^ of the eight articles addressing effects of BDNF polymorphism had available FM scores assessed at the end of the intervention to be used for a meta-analysis. Two studies^59,60^ also included a retention assessment, with that data being included for a second meta-analysis.

### BDNF val^66^met polymorphism

In total, 598 individuals (59.2% men, 40.8% women) sustaining a stroke participated in the eight studies included in the qualitative analysis. The average age of the participants (mean ± SD) was 58.4 ± 3.2 years A greater proportion of participants had sustained ischemic strokes (79.7%) compared to hemorrhagic strokes (20.3%). The distribution of the more-affected side was almost equal (50.7% right, 49.3% left). Four^57,61–63^ of the included studies were ranked as *‘good’* and the reaming six^23,58–60,64,65^ *‘fair’*(*Supplementary Table 1*). Participants were either in the acute^57–60,64^ or chronic^23,61^ stage post-stroke. All participants had moderate-to-severe^66^ UL motor impairment (FM score ≤49/66).

Table 1 presents a summary of studies evaluating the effects of BDNF polymorphism with a focus on sample size, type and dose of rehabilitation provided (if any), main outcomes and results. The sample size used for the Meta-analysis was 295 (no polymorphism: 101, polymorphism: 194). Analysis revealed a *large* (0.50, 95% CI: 0.11 - 0.88, p = 0.01, I^2^ = 54%, random effects model; Figure 2) effect size at the end of the intervention period for improvement in UL FM scores in those without compared to those with the polymorphism. At retention testing, the sample size used was 79 (no polymorphism: 19, polymorphism: 60,). We found a similar large effect size (0.58, 95% CI: 0.06 - 1.11, p = 0.03, I^2^ = 0%, random effects model; Figure 3).

**Figure 2.**
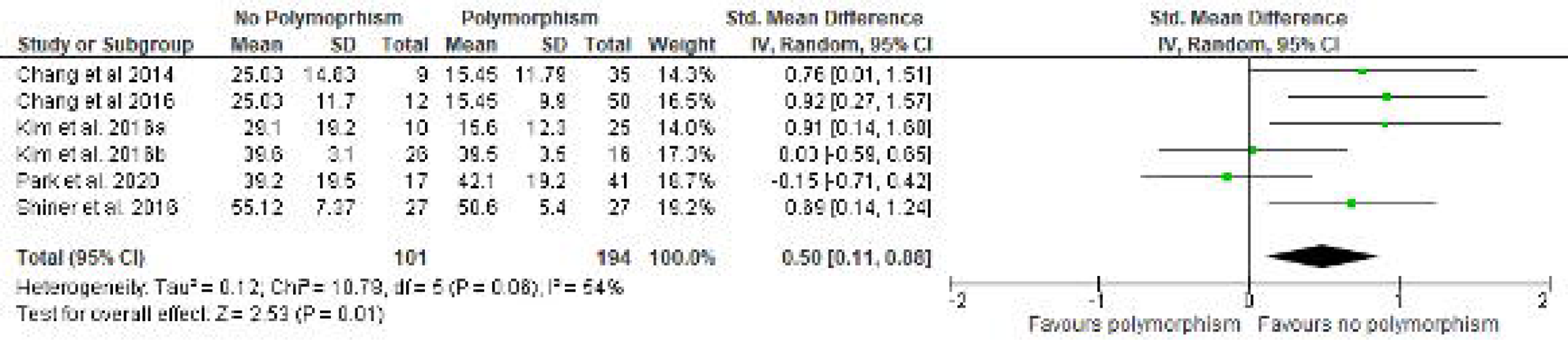
Results of meta-analyses examining influence of genetic polymorphisms on upper limb motor impairment quantified using the Fugl-Meyer Assessment, at the end of the intervention period. Larger squares indicate bigger study effect sizes. The diamonds represent pooled effects of results of individual studies. The location of the diamond indicates the estimated effect size and precision of the estimate is indicated by the width of the diamond

**Figure 3.**
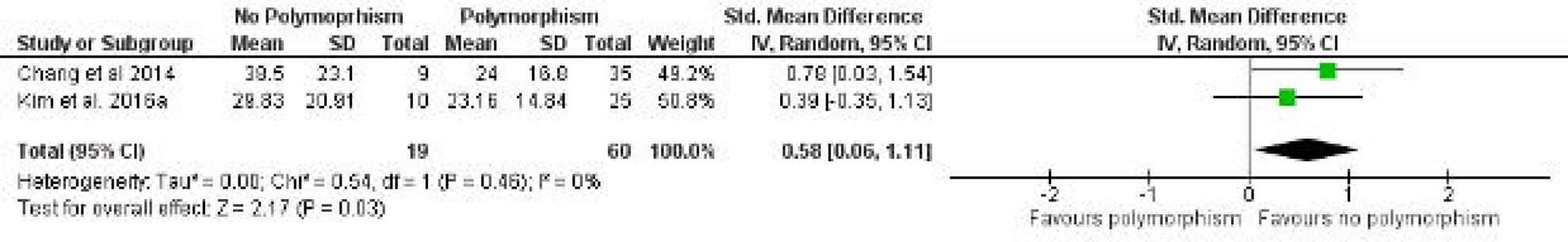
Results of meta-analyses examining influence of genetic polymorphisms on upper limb motor impairment quantified using the Fugl-Meyer Assessment, at retention testing. Larger squares indicate bigger study effect sizes. The diamonds represent pooled effects of results of individual studies. The location of the diamond indicates the estimated effect size and precision of the estimate is indicated by the width of the diamond.

**Table 1.** Effect of BDNF polymorphism.

| Study; Sample size (n);<br>Val/Val and Met Allele<br>distribution and<br>Down's and Black<br>score | Intervention | Rehabilitation provided/<br>Dose | Outcomes and timing of<br>assessment | Results |
| --- | --- | --- | --- | --- |
| Chang et al. 2014;<br>n = 44;<br>Val/Val: n=9;<br>Met allele: n = 35<br>DBS: 19 (Good) | 10 sessions of rTMS<br>over two weeks.<br>Each session had 50<br>trains of 10 Hz<br>frequency for 5<br>seconds at 90% RMT | Each train of rTMS was<br>followed by 50 seconds of<br>reaching and grasping<br>exercises.<br>Active and active assisted<br>exercises consisting of range<br>of motion exercises, moving,<br>and grasping and releasing<br>cups and cubes.<br>All participants also received<br>conventional physical (PT)<br>and occupational therapy<br>(OT) sessions, involving gait,<br>fitness, and ADL training for<br>3 hours each day. | • Upper and lower limb<br>Fugl-Meyer<br>Assessment (FMA)<br>scores<br>• Box and Blocks test<br>(BBT).<br>Assessments conducted at<br>baseline, end of the<br>intervention and 2-month<br>retention. | <u>FMA</u><br>• UL: Greater change seen in Val/Val<br>group at post (10 points) and<br>retention (23 points) compared to<br>Met alleles (4 and 11 points)<br>respectively (p<0.05).<br>• LL: Both groups improved at both<br>assessments with no between group<br>difference.<br><u>BBT:</u><br>• Greater change seen in Val/Val<br>group (16 blocks) compared to Met<br>alleles (6 blocks; p<0.05) at<br>retention. |
| Chang et al 2016;<br>n = 62;<br>Val/Val: n=12<br>Met allele; n= 50<br>DBS: 18 (fair) | 10 sessions of rTMS<br>over two weeks.<br>Each session had 20<br>trains of 50 stimuli<br>of 10 Hz frequency<br>at 90% RMT | Each train of rTMS was<br>followed by 50 seconds of<br>reaching and grasping<br>exercises.<br>All participants also received<br>conventional PT and OT<br>sessions, involving gait,<br>fitness, and ADL training for<br>3 hours each day. | • Upper and lower limb<br>and total FMA scores<br>• Degree of preserved<br>Corticospinal tract<br>(CST) integrity<br>quantified by<br>Diffusion tensor<br>imaging and<br>presence/absence of<br>MEP in the FDI<br>muscle. | • 20 participants were good<br>responders and 42 were poor<br>responders.<br>• Greater proportion of good<br>responders had Val/Val genotype<br>(35%) compared to poor responders<br>(11%).<br>• Those with Val/Val genotype had<br>significantly higher change in UL<br>FMA scores (13.7 points) compared<br>to Met alleles (1 point) |
|  |  |  | Assessments conducted at baseline and end of the intervention.<br>Patients classified as good or poor responders depending upon the amount of change in UL FMA scores: good responders ($\geq 5$ points); poor responders ($\leq 4$ points) | • Individuals with Val/Val genotype almost twice more likely to have better improvement than those with Met alleles. |
| Kim et al. 2016a<br>n = 35;<br>Val/Val: n=10<br>Met allele; n= 25<br>DBS: 17 (Fair) | No details provided | No information provided | • UL FMA scores<br>• Values of fractional anisotropy (FA), axial diffusivity (AD) and radial diffusivity (RD) for CST.<br>Assessments conducted at baseline, T1 (1 month after baseline) and T3 (3 months after baseline) | *UL FMA scores*<br>• Greater change in the *Val/Val* group at T2 (7 points) and T3 (17 points) compared to the *Met* group (4 and 12 points respectively).<br>• Moderate positive correlation with FA Values at T1 ($r = 0.78$) and T2 ($r = 0.72$) for the *Val/Val* group and at T3 ($r = 0.59$) for the *Met* group<br>• Moderate positive correlation with AD scores at T1 ($r = 0.78$) and T2 ($r = 0.72$) for the *Val/Val* group.<br>• Moderate negative correlation with RD Values scores at T3 ($r = -0.59$) for the *Met* group. |
| Kim et al. 2016b<br>n = 42;<br>Val/Val: n=26<br>Met allele; n= 16<br>DBS: 16 (Fair) | Robotic therapy spread over 2-3 weeks. | Therapy consisted of repeated grasp and release movements of the affected hand and wrist.<br>Participants wore a Hand Wrist Assistive Rehabilitation Device and practiced tasks with real objects as well as virtual reality games (e.g., | • UL FMA scores<br>• Values of percentage change on fMRI signals and activation volume obtained from the ipsi- and contralesional primary sensorimotor cortex | • No difference in change in UL FMA scores between the *Val/Val* group at T2 (2.1 points) and the *Met* group (3.2 points) at the end of therapy.<br>• However, the *Val/Val* group had greater percentage signal change ($p = 0.037$) and activation volume ($p = 0.03$) in the ipsilesional primary |
|  |  | squeezing lemons, moving jewels into a safe, etc.)<br>Tasks focusing on speed, reaction time, force, and range of motion were also practiced. | and dorsal premotor cortex using fMRI.<br>Assessments conducted at baseline, T1 (end of therapy) | sensorimotor cortex compared to the those in the Met group. |
| Shiner et al. 2016<br>n = 54;<br>Val/Val: n=27<br>Met allele: n=27<br>DBS: 19 (Fair) | Wii based movement therapy (n=40) or modified constraint induced movement therapy (mCIMT, n=14). | 10 one-hour long sessions on consecutive weekdays.<br>Sessions targeted movements of the more-affected hand and arm.<br>The Wii group played golf, bowling, baseball, boxing, and tennis using the controller in the more-affected arm.<br>mCIMT group received task-based training on object manipulation focusing on movement speed.<br>Individuals classified into those with low (inability to move >1 block on BBT), moderate (inability to complete Perdue Pegboard Test) and high (ability to complete Perdue Pegboard Test) functional ability. | • Upper limb FMA<br>• Wolf Motor Function Test - timed task (WMFT- tt)<br>• Motor Activity Log Quality of Movement (MAL-QoM) scores.<br>Assessments conducted at baseline and at the end of the intervention. | • All participants improved on FMA, WMFT -TT and MALQoM scores at the end of the interventions.<br>• Overall, no difference in amount of change in FMA, WMFT-tt and MAL-QoM scores between those with Val/Val and Met alleles.<br>• However, subgroup analysis revealed *less change* in those with Met alleles and moderate (8.9%) or high (13.8%) functional ability on WMFT-tt scores compared to those with Val/Val (25.2% and 37.3% respectively).<br>• Similar results were seen on FMA Values *less change* in those with Met alleles and moderate (7%) or high (2%) functional ability compared to those with Val/Val (10% and 4% respectively). |
| Chang et al. 2017;<br>n = 97;<br>Val/Val: n=21<br>Met allele: n=76 | Traditional inpatient rehabilitation | Traditional inpatient (2 hrs PT, 1 hr OT) followed by outpatient rehab (1 hr PT, 30 mins OT) or home exs. | • Upper limb FMA scores.<br>Assessments conducted at baseline and T1 (after 3 months). | *Individuals with normal FMA scores or mild and moderate impairment* |
| DBS: 14 (Fair) |  |  | Participants classified into 4 categories based upon FMA scores at T1: normal: 66, mild impairment (41–65), moderate impairment (25–40), and severe 0–24). | • Baseline FMA scores explained 47% of variance in FMA scores at T1<br>*Individuals with severe motor impairment*<br>• A combination of presence of Met alleles, baseline FMA scores and age explained 59.5% of the variance in FMA scores at T1. Individuals with Met alleles were *1.48 times less likely* to have better scores on the UL FMA.<br>• Smaller proportion of individuals with two (10%) or one Met allele (31%) recovered significantly at T1 compared to those with Val/Val genotype (42.9%).<br>• Significant correlation between number of Met alleles and FMA score at T1 ($\rho = -0.248$, $p < 0.05$). |
| Park et al. 2020;<br>DBS: n = 58;<br>Val/Val: n=17<br>Met allele: n=41<br>17 (Fair) | Traditional inpatient rehabilitation | All participants received the same dose of PT and OT (3-week intensive inpatient rehabilitation). | • Upper limb FMA scores.<br>• FA Values for CST, intrahemispheric connection from M1 to the ventral premotor cortex and corpus callosum (CC).<br>Assessments conducted at baseline and T2 (after 3 months). | *UL FMA scores*<br>• Similar mean change seen in UL FMA scores in those with Val/Val (12.6 points) and Met alleles (13.8 points) at T2.<br>• *In the Val/Val group*, moderate negative correlation with FA in contralesional intrahemispheric connection from M1 to the ventral premotor cortex at T2 ($r = -0.60$; $p = 0.024$). |
|  |  |  |  | • *In those with Met alleles*, moderate positive correlation with FA in the in the ipsilesional CST ($r=0.47$; $p=0.003$) and FA in the CC ($r=0.41$, $p=0.011$). |
| Cramer et al 2022;<br>n = 206;<br>Val/Val: n = 166;<br>Met allele: n = 40<br>DBS: 21 (Good) | Task Oriented upper extremity training or OT | Participants were randomized to 30 hrs each of task-oriented upper extremity training (Accelerated Skill Acquisition Program), dose-equivalent occupational therapy, or standard of care. | • Change in Log WMFT-tt.<br>• Cerebral atrophy measured using ventricular brain ratio.<br>Assessments carried out at baseline and at end of 12 months | • Overall, no difference in amount of change in Log WMFT-tt scores between individuals with Met alleles compared to those with Val/Val genotype.<br>• Greater cerebral atrophy ($p<0.01$) seen in individuals with Met alleles compared to those with Val/Val genotype.<br>• This enlargement was caused primarily by an enlargement in ventricular volume ($p=0.0098$). |

*ADL: Activities of Daily Living ;DBS: Downs and Black Checklist Score; FDI: Flexor Digitorum Indicis; fMRI: Functional Magnetic Resonance Imaging; MEP: Motor Evoked Potential; rTMS: Repetitive Transcranial Magnetic Stimulation; RMT: Resting Motor Threshold.*

In addition to UL FM scores, other assessments at the body structure and function level included use of functional magnetic resonance imaging (fMRI), diffusion tensor imaging (DTI) and MRI. In terms of fMRI outcomes, lower ipsilesional activation volume and percentage signal change were noted in individuals with the Met alleles as compared to the Val homozygous individuals.^23^ Use of DTI revealed differences in radial and axial diffusion^59^ and fractional anisotropy^60^ between individuals with and without met alleles. Individuals with met alleles also had greater cerebral atrophy on MRI.^62^

Sensitivity analysis included an additional meta-analysis being conducted with data from five studies included in this analysis. The only study excluded^59^ provided no details on whether and if so, how many sessions of any form of rehabilitation were provided to the participants. The sample size used for this Meta-analysis was 260 (no polymorphism: 91, polymorphism:169). Analysis revealed a large (0.43, 95% CI: 0.01 - 0.86, p = 0.046, I^2^ = 57%, random effects model; Figure 4) effect size at the end of the intervention period for improvement in UL FM scores in those without compared to those with the polymorphism.

**Figure 4.**
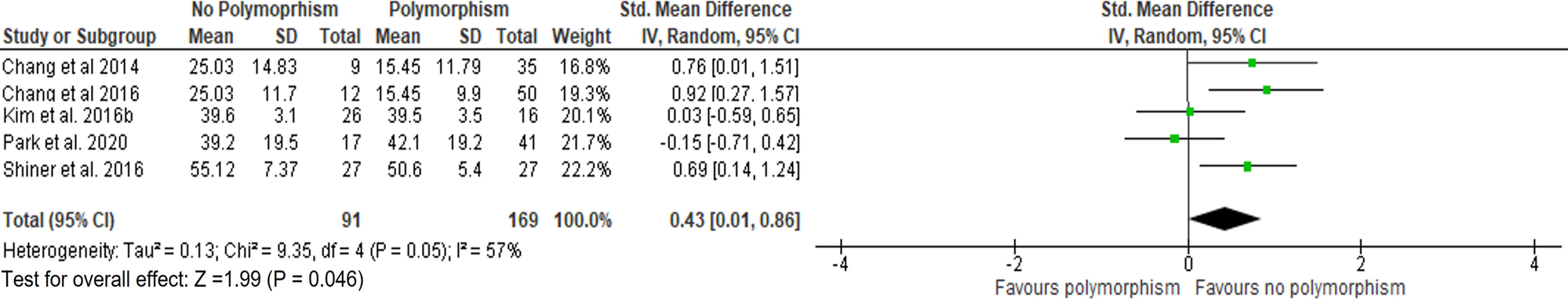
Results of sensitivity analysis (meta-analyses) examining influence of genetic polymorphisms on upper limb motor impairment quantified using the Fugl-Meyer Assessment, at the end of the intervention. Larger squares indicate bigger study effect sizes. The diamonds represent pooled effects of results of individual studies. The location of the diamond indicates the estimated effect size and precision of the estimate is indicated by the width of the diamond.

#### APOEɛ4 Polymorphism

Table 2 presents a summary of studies evaluating the effects of APOEɛ4 and COMT val^158^met polymorphism. Two (good quality^61,62^) of the eight studies examining the effects of BDNF val66met polymorphism also assessed the effects of APO ɛ4 polymorphism. These two studies included a total of 260 participants (61.5% men, 38.5% women). A greater proportion of participants had sustained ischemic strokes (83.4%) compared to hemorrhagic strokes (16.7%). The distribution of the more-affected side was equal (50 % right, 50% left).

**Table 2.** Effect of APOE and COMT polymorphism.

| Study; Sample size (n);<br>distribution and<br>Down's and Black<br>score | Intervention | Rehabilitation provided/<br>Dose | Outcomes and timing of<br>assessment | Results |
| --- | --- | --- | --- | --- |
| A. Effects of APOE polymorphism |  |  |  |  |
| Shiner et al. 2016;<br>n = 54;<br>$\epsilon 4$: n = 9;<br>non $\epsilon 4$: n = 45<br>DBS: 19 (Good) | Wii based movement<br>therapy (n=40) or<br>modified constraint<br>induced movement<br>therapy (mCIMT,<br>n=14). | 10 one-hour long sessions on<br>consecutive weekdays.<br>Sessions targeted movements<br>of the more-affected hand and<br>arm.<br>The Wii group played golf,<br>bowling, baseball, boxing, and<br>tennis using the controller in<br>the more-affected arm.<br>mCIMT group received task-based training on object<br>manipulation focusing on<br>movement speed.<br>Individuals classified into those<br>with low (inability to move >1<br>block on BBT), moderate<br>(inability to complete Perdue<br>Pegboard Test) and high<br>(ability to complete Perdue<br>Pegboard Test) functional<br>ability. | • Upper limb FMA<br>• Wolf Motor Function Test - timed task (WMFT- tt)<br>• Motor Activity Log Quality of Movement (MAL-QoM) scores.<br>Assessments conducted at baseline and post-intervention. | • All participants improved on FMA, WMFT-TT and MALQoM scores at the end of the interventions.<br>• Overall, no difference in amount of change in FMA and MAL-QoM scores between $\epsilon 4$ carriers and those with non $\epsilon 4$ genotype.<br>• $\epsilon 4$ carriers tended to take longer to complete ADL activities (WMFT-tt, p = 0.057) compared to those with non $\epsilon 4$ genotype. |
| Cramer et al 2022;<br>n = 206;<br>$\epsilon 4$: n = 61;<br>non $\epsilon 4$: n = 145<br>DBS: 21 (Good) | Task Oriented upper<br>extremity training or<br>Occupational therapy | Patients were randomized to<br>30 h each of task-oriented<br>upper extremity training<br>(Accelerated Skill Acquisition<br>Program), dose-equivalent<br>occupational therapy, or<br>standard of care. | • Change in Log WMFT-tt.<br>• Cerebral atrophy measured using ventricular brain ratio. | • Overall, no difference in amount of change in Log WMFT-tt scores between $\epsilon 4$ carriers and those with non $\epsilon 4$ genotype.<br>• No differences seen in cerebral atrophy between individuals with and without the polymorphism. |
|  |  |  | Assessments carried out at baseline and at end of 12 months. |  |
| *B. Effects of COMT polymorphism* |  |  |  |  |
| Liepert et al. 2013;<br>n = 83,<br>val/val = 12,<br>met allele: 71<br>DBS: 19 (Good) | Traditional Rehabilitation | Rehabilitation program included PT, OT, endurance, and strength training.<br>Program was adapted to individual needs of the patient.<br>Details unavailable on total duration of therapy. | Rivermead Motor Assessment (RMA) and Barthel Index (BI) scores. RMA scores divided into Gross Function, Leg and trunk and Upper limb function.<br>Assessments carried out at baseline, 4 weeks later and at the end of 6 months. | *RMA scores*<br>Individuals with two met alleles showed less improvement in *gross function* ($p = 0.003$), *leg and trunk function* ($p = 0.022$) as well as *upper limb function* ($p = 0.047$).<br>Significant correlation with BI scores at all time points ($p < 0.001$). |
| Kim et al. 2016<br>n = 74,<br>val/val = 41,<br>met allele = 33<br>DBS = 15 (fair) | No details provided | No details provided. | Total Fugl-Meyer Assessment (FMA) and Functional Independence Measure (FIM) total scores.<br>Assessments carried out at hospital admission, discharge 3- and 6-mos. post-discharge. | *FMA scores*<br>Lower scores at discharge, 3-mos and 6 mos post discharge assessments in those with met alleles ($p < 0.01$) compared to the Val heterozygous group.<br>*FIM scores*<br>Lower scores at discharge ($p < 0.01$), 3-mos and 6 mos post discharge ($p < 0.05$) assessments in those with met alleles compared to the Val heterozygous group. |

DBS: Downs and Black Checklist Score; OT: Occupational Therapy; PT: Physical Therapy;

Both studies used the Wolf Motor Function Test - timed test (WMFT-tt) as the primary outcome. No differences were noted between individuals with and without the polymorphism on WMFT-tt scores (Table 2A). In addition, groups did not differ on the amount of change seen in UL FM scores and self-reported levels of UL quality (assessed using the Motor Activity Log)^61^ or in the amount of cerebral atrophy noted between groups.^62^

#### COMT val^158^met Polymorphism

Two fair^63,65^ quality studies including 157 participants (59.7% men, 40.3% women) examined the influence of COMT val^158^met polymorphism (53: no polymorphism, 104: polymorphism). A greater proportion of participants had sustained ischemic strokes (83.8%) compared to hemorrhagic strokes (16.2%). The distribution of the more-affected side was 46.8% right side, 51.9 %, left side and 1.3% of the participants had bilateral strokes. The studies used either the UL section of the Rivermead Motor Assessment (RMA)^63^ or the FMA.^65^

Compared to those with met^158^met, participants with val^158^met allele (ES = 0.51) or val^158^val (ES = 0.76) distribution had greater recovery with large effect sizes on the UL section of the RMA, at the end of the intervention period. Similar results were obtained for the FMA. Individuals with val^158^val distribution had greater recovery on the FMA at the end of the intervention period (ES = 2.69), and at 3 (ES = 1.51) and 6 months (ES = 1.98) retention testing. In addition, participants without the polymorphism improved more on other components of the RMA scores^63^ including gross function, leg, and trunk function and higher FIM scores.^65^

### Risk of Bias

Overall, the risk of bias was low for all studies (*Supplementary Figure 1, Supplementary Table 2*), except one.^59^ The ROB for this one study could not be ascertained for the domains of measurement of interventions and departures from intended interventions, as information on whether the participants received any intervention or not was missing.

## Discussion

Results from this systematic review and meta-analysis showed that the presence of some genetic polymorphisms negatively influence post-stroke UL motor improvement. The meta-analyses revealed that BDNF polymorphism negatively impacted UL motor improvement, immediately after the end of the intervention period as well as at retention testing. Overall, majority of the studies had low risk of bias, which lends further credence to these results. Sensitivity analyses revealed that results continued to remain significant even with the exclusion of the study where information on some domains of bias was not available.

These results are in agreement to those found previously,^42^ and extend those findings more specifically to UL motor improvement and not just general recovery from a stroke. We also found that while APO ɛ4 polymorphism does not influence UL motor improvement, presence of COMT val^158^met polymorphism has a negative impact. Our results for APO ɛ4 agree partially with those found previously,^39^ and go beyond those results by focusing on UL motor improvement. To our knowledge, this is the first review which has systematically investigated the effects of COMT polymorphism on post-stroke UL motor improvement.

### Study Quality Assessment

Of the ten articles included in the review, four^57, 61–63^ were ranked as *‘good’* and the reaming six^23, 58–60,64,65^ *‘fair’.* We did not find any articles that could be categorized as being of ‘*poor’ quality*. However, we also did not have any articles that were ‘*excellent’* in quality. We used the modified D&B checklist in this review, as both randomized and non-randomized study designs were included. The modified D&B checklist score includes an assessment of internal and external validity, reporting standards and sample size. Commonly non-reported details across studies in this review include information on external validity (three questions) and on power/sample size analysis. Inherent word limitations in manuscript length may often preclude exclusion of such information in the main text. It is suggested that such information may be reported at least as supplementary material to provide a better overview of the rationale behind participant selection in the studies.

### Interventions used and number of sessions

A variety of interventions were used amongst the various studies included in the review. The interventions used included the use of rTMS along with traditional physical (PT) and occupational therapy (OT) sessions,^57,58^ provision of traditional PT and OT sessions alone^60,63, 64^ virtual reality platform along with robotic assistive devices,^23^ commercial gaming solution (i.e., Nintendo Wii),^61^ modified constraint induced movement therapy,^61^ and task-oriented UL training.^62^ No details were provided for two studies.^59,65^ The above-mentioned interventions were delivered at different intensities. Time spent in therapy was the most common metric used to denote intensity in the included studies. Time spent in therapy was either 60 minutes/day,^61,62^ 90 minutes/day (outpatient rehabilitation phase)^64^ or three hours/day.^57,58,64^ Therapy was provided for 10 sessions over 2 weeks,^57,58, 61^ 2-3 weeks^23,60^ or 30 sessions.^62^ Information on exact number of sessions was not provided for the other studies.

Although time spent in therapy is one metric of intensity,^67^ other metrics include numbers of repetitions^68^ as well as “*amount of physical and/or mental work put forth by the client”*.^69^ Previous work involving healthy controls with BDNF polymorphism has shown that employing a high number of repetitions (about 800 repetitions/session) for about five days can cause significant changes in short-term plasticity even in those with the polymorphism.^70^ The minimal number of repetitions/ session in individuals with BDNF and other polymorphisms that have also sustained a stroke are currently unknown. Approaches similar to those used previously could be employed to estimate the minimal number of repetitions to achieve a plateau in motor performance in a single session.^71,72^ Whether using a fixed number of repetitions results in better UL motor improvement in post-stroke individuals with polymorphisms remains to be estimated.

### Outcomes used to assess improvement

A variety of outcomes were used to assess the effects of the different polymorphisms. At the body structure and function level, in addition to the FMA, the Rivermead Motor Assessment, MRI, fMRI and diffusion tensor imaging (DTI) were used. At the activity level, Box and Blocks Test, Wolf Motor Function Test and Motor Activity Log helped specifically assess UL activity performance, while outcomes including Barthel Index and Functional Independence measure helped assess general activity performance. All the selected outcomes have well established psychometric properties^73,74^ and measures including the FMA, Functional Independence Measure, Motor Activity Log, Rivermead Motor Assessment, and Wolf Motor Function Test are amongst recommended measures.^73^ However, no study used any assessment at the participation level. Hence the effects of the polymorphism at the participation level have not yet been assessed. If select core measures such as those recommended by previous publications^75,76^ are used, the effects across the different levels of the ICF could be better understood. In addition, the UL part of the FM does not account for the use of altered movement patterns.^66^ It is currently unknown whether individuals with genetic polymorphisms use compensatory movement patterns for task completion.

### Influence of ethnicity

Majority of the studies in this review emerged from Asia, particularly from South East Asia, with only four studies^23,61–63^ being conducted outside Asia. Amongst these four studies, three^23,61, 62^ had detailed demographics available on ethnicity of the participants. Individuals belonging to Asian Ethnicity already have poor outcomes after a stroke.^77^ There are some reports that individuals of Asian Ethnicity tend of receive less rehabilitation services compared to individuals from a Caucasian ethnicity and have higher rates of hospital readmission.^78–80^ In addition, in all the three biomarkers examined in this meta-analysis, individuals with Asian ethnicity have higher rates of polymorphism.^81–83^ The presence of high rates of the polymorphism can be an additional factor explaining the lower rates of post-stroke motor improvement seen in this population. This information can likely play an important role in prediction of prognosis after a stroke. Furthermore, it can also help make decisions as to whether and if so, the extent to which provision of rehabilitation interventions need to differ for this population.

## Limitations

We only included studies involving adult participants published in English (since no one in the team was proficient in other languages). It might be possible that we missed studies published in other languages. None of the studies had an explicit sample size calculation. Information on baseline levels of depression and/or intake of anti-depression medication was available in only two studies.^23, 63^ Information on the presence of depression is essential, as the presence of genetic polymorphisms is an additional risk factor^84^ for post-stroke depression and can influence the extent of UL motor improvement.^10^

## Conclusion

Our review and meta-analyses results indicate that presence of genetic polymorphisms in BDNF and COMT negatively impact post-stroke motor improvement. This is especially true at the body-structure and function domain of the ICF. Our findings may contribute to the understanding of one of the underlying mechanisms to help explain some variability in post-stroke UL motor improvement. This is valuable information for the means of tailoring a plan of care, creating realistic goals, and providing relevant, individualized care to every patient. In addition, new questions have been identified including does the i) use of a fixed number of repetitions result in similar or better levels of UL motor improvement in individuals with genetic polymorphisms; ii) presence of COMT val^158^met continue to influence motor improvement at retention testing; iii) presence of genetic polymorphisms influence participation levels and iv) do individuals with genetic polymorphisms use altered movement patterns and if so, to what extent. Answers to these emergent questions can help better understand the influence of genetic polymorphisms on post-stroke upper limb motor improvement.

## Data Availability

All data produced in the present work are contained in the manuscript

## Declarations

### Ethics approval and consent to participate

Not applicable, as this manuscript is a systematic review. However, ethics approval was obtained for all the studies included.

### Consent for publication

Not applicable

### Author contributions

**Sandeep K Subramanian:** Conceptualization; Funding acquisition; Methodology; Project administration; Formal Analysis, Supervision; Writing – review & editing.

**Riley T Morgan:** Investigation, Formal Analysis, Data Curation, Writing – Original draft.

**Carl Rasmusson:** Investigation, Formal Analysis, Data Curation, Writing – Original draft.

**Kayla M Shepherd:** Investigation, Formal Analysis, Data Curation, Writing – Original draft.

**Carol L Li:** Conceptualization; Methodology; Writing – review & editing.

## Acknowledgments

The authors would like to acknowledge the UT Health San Antonio Briscoe library for access to numerous databases.

## Availability of data and materials

Not applicable

## References

1. Feigin VL, Vos T, Alahdab F, et al. Burden of Neurological Disorders Across the US From 1990-2017: A Global Burden of Disease Study. JAMA Neurol 2021; 78: 165–176.

2. Winstein CJ, Stein J, Arena R, et al. Guidelines for adult stroke rehabilitation and recovery - A guideline for healthcare professionals from the American Heart Association/American Stroke Association. Stroke 2016; 47: e98–e169.

3. Cassidy JM, Cramer SC. Spontaneous and therapeutic-induced mechanisms of functional recovery after stroke. Transl Stroke Res 2017; 8: 33–46.

4. Buma F, Kwakkel G, Ramsey N. Understanding upper limb recovery after stroke. Restor Neurol Neurosci 2013; 31: 707–722

5. Newell KM. Motor skill acquisition. Annu Rev Psychol 1991; 42: 213–237.

6. Subramanian SK, Lourenco CB, Chilingaryan G, et al. Arm motor recovery using a virtual reality intervention in chronic stroke: randomized control trial. Neurorehabil Neural Repair 2013; 27: 13–23.

7. White JH, Bartley E, Janssen H, et al. Exploring stroke survivor experience of participation in an enriched environment: a qualitative study. Disabil Rehabil 2015; 37: 593–600.

8. Kleim JA. Neural plasticity and neurorehabilitation: teaching the new brain old tricks. J Commun Disord 2011; 44: 521–528.

9. Levy T, Christie LJ, Killington M, et al. “*Just that four letter word, hope*”: stroke survivors’ perspectives of participation in an intensive upper limb exercise program; a qualitative exploration. Physiother Theory Pract 2021: 1–15.

10. Subramanian SK, Chilingaryan G, Sveistrup H, et al. Depressive symptoms influence use of feedback for motor learning and recovery in chronic stroke. Restor Neurol Neurosci 2015; 33: 727–740.

11. Stewart JC, Cramer SC. Genetic variation and neuroplasticity: Role in rehabilitation after stroke. J Neurol Phys Ther 2017; 41 Suppl 3: S17–S23.

12. Bernhardt J, Borschmann K, Boyd L, et al. Moving rehabilitation research forward: Developing consensus statements for rehabilitation and recovery research. Int J Stroke 2016; 11: 454–458.

13. Feng W, Wang J, Chhatbar PY, et al. Corticospinal tract lesion load: an imaging biomarker for stroke motor outcomes. Ann Neurol. 2015; 78: 860–870.

14. Connell LA, Smith MC, Byblow WD, et al. Implementing biomarkers to predict motor recovery after stroke. NeuroRehabilitation 2018; 43: 41–50.

15. Musunuru K, Hickey KT, Al-Khatib SM, et al. Basic concepts and potential applications of genetics and genomics for cardiovascular and stroke clinicians: a scientific statement from the American Heart Association. Circ. Cardiovasc. Genet 2015; 8: 216–242.

16. Pearson-Fuhrhop KM, Kleim JA, Cramer SC. Brain plasticity and genetic factors. Top Stroke Rehabil 2009; 16: 282–299.

17. Uhm KE, Kim YH, Yoon KJ, et al. BDNF genotype influence the efficacy of rTMS in stroke patients. Neurosci Lett 2015; 594: 117–121

18. Langdon KD, Corbett D. Improved working memory following novel combinations of physical and cognitive activity. Neurorehabil Neural Repair 2012; 26: 523–532.

19. Mang CS, Campbell KL, Ross CJ, et al. Promoting neuroplasticity for motor rehabilitation after stroke: considering the effects of aerobic exercise and genetic variation on brain-derived neurotrophic factor. Phys Ther 2013; 93: 1707–1716.

20. Casey BJ, Glatt CE, Tottenham N, et al. Brain-derived neurotrophic factor as a model system for examining gene by environment interactions across development. Neuroscience 2009; 164: 108–120.

21. Egan MF, Kojima M, Callicott JH, et al. The BDNF val66met polymorphism affects activity-dependent secretion of BDNF and human memory and hippocampal function. Cell 2003; 112: 257–269.

22. McHughen SA, Rodriguez PF, Kleim JA, et al. BDNF val66met polymorphism influences motor system function in the human brain. Cereb Cortex 2010; 20: 1254–1262.

23. Kim DY, Quinlan EB, Gramer R, et al. BDNF Val66Met Polymorphism is related to motor system function after stroke. Phys Ther 2016; 96: 533–539.

24. Balkaya M, Cho S. Genetics of stroke recovery: BDNF val66met polymorphism in stroke recovery and its interaction with aging. Neurobiol Dis 2019; 126: 36–46.

25. Meyer-Lindenberg A, Kohn PD, Kolachana B, et al. Midbrain dopamine and prefrontal function in humans: interaction and modulation by COMT genotype. Nat Neurosci 2005; 8: 594–596.

26. Floel A, Hummel F, Breitenstein C, et al. Dopaminergic effects on encoding of a motor memory in chronic stroke. Neurology 2005; 65: 472–474.

27. Cramer SC. Drugs to enhance motor recovery after stroke. Stroke 2015; 46: 2998–3005.

28. Lachman HM, Papolos DF, Saito T, et al. Human catechol-O-methyltransferase pharmacogenetics: description of a functional polymorphism and its potential application to neuropsychiatric disorders. Pharmacogenetics 1996; 6: 243–250.

29. Egan MF, Goldberg TE, Kolachana BS, et al. Effect of COMT Val108/158 Met genotype on frontal lobe function and risk for schizophrenia. Proc Natl Acad Sci U S A 2001; 98: 6917–6922.

30. Pearson-Fuhrhop KM, Minton B, Acevedo D, et al. Genetic variation in the human brain dopamine system influences motor learning and its modulation by L-Dopa. PLoS One 2013; 8: e61197. DOI: 10.1371/journal.pone.0061197.

31. Yin Y, Liu Y, Xu M, et al. Association of COMT rs4680 and MAO-B rs1799836 polymorphisms with levodopa-induced dyskinesia in Parkinson’s disease-a meta-analysis. Neurol Sci 2021. DOI: 10.1007/s10072-021-05509-3.

32. Galderisi S, Maj M, Kirkpatrick B, et al. Catechol-O-methyltransferase Val158Met polymorphism in schizophrenia: associations with cognitive and motor impairment. Neuropsychobiology 2005; 52: 83–89.

33. Doyon J, Bellec P, Amsel R, et al. Contributions of the basal ganglia and functionally related brain structures to motor learning. Behav Brain Res 2009; 199: 61–75.

34. Huotari M, Gogos JA, Karayiorgou M, et al. Brain catecholamine metabolism in catechol-O-methyltransferase (COMT)-deficient mice. Eur J Neurosci 2002; 15: 246–256.

35. Parasuraman R, Greenwood PM, Sunderland T. The apolipoprotein E gene, attention, and brain function. Neuropsychology 2002; 16: 254–274.

36. McFadyen CA, Zeiler FA, Newcombe V, et al. Apolipoprotein E4 polymorphism and outcomes from traumatic brain injury: a living systematic review and meta-analysis. J Neurotrauma 2021; 38: 1124–1136.

37. Strattan LE, Britsch DRS, Calulot CM, et al. Novel influences of sex and APOE genotype on spinal plasticity and recovery of function after spinal cord injury. eNeuro 2021; 8. DOI: 10.1523/ENEURO.0464-20.2021.

38. Martinez-Gonzalez NA, Sudlow CL. Effects of apolipoprotein E genotype on outcome after ischaemic stroke, intracerebral haemorrhage and subarachnoid haemorrhage. J Neurol Neurosrg Psychiatr 2006; 77: 1329–1335.

39. Math N, Han TS, Lubomirova I, et al. Influences of genetic variants on stroke recovery: a meta-analysis of the 31,895 cases. Neurol Sci 2019; 40: 2437–2445.

40. Levin MF, Kleim JA, Wolf SL. What do motor “recovery” and “compensation” mean in patients following stroke? Neurorehabil Neural Repair 2009; 23: 313–319.

41. Subramanian SK, Chilingaryan G, Sveistrup H, et al. Influence of training environment and cognitive deficits on use of feedback for motor learning in chronic stroke. In: Deutsch J, Wright WG, Weiss PT, eds. Proceedings of the 2015 International Conference on Virtual Rehabilitation (ICVR); June 9–12, 2015; Valencia, Spain. IEEE Publications. doi: 10.1109/ICVR.2015.7358582

42. Liu X, Fang JC, Zhi XY, et al. The influence of val66met polymorphism in brain-derived neurotrophic factor on stroke recovery outcome: A systematic review and meta-analysis. Neurorehabil Neural Repair 2021; 35: 550–560.

43. Kotlega D, Peda B, Zembron-Lacny A, et al. The role of brain-derived neurotrophic factor and its single nucleotide polymorphisms in stroke patients. Neurol Neurochir Pol 2017; 51: 240–246.

44. Veerbeek JM, van Wegen E, van Peppen R, et al. What is the evidence for physical therapy poststroke? A systematic review and meta-analysis. PloS one 2014; 9: e87987.

45. Subramanian SK, Birnbaum LA. Genetic polymorphisms negatively impact upper limb motor recovery after stroke-A meta-analysis [abstract]. Stroke 2019; 50(Suppl_1): AWP193.

46. Morton S, Barton CJ, Rice S, et al. Risk factors and successful interventions for cricket-related low back pain: a systematic review. Br J Sports Med 2014; 48: 685–691.

47. Downs SH, Black N. The feasibility of creating a checklist for the assessment of the methodological quality both of randomised and non-randomised studies of health care interventions. J Epidemiol Comm Health 1998; 52: 377–384.

48. Subramanian SK, Caramba SM, Hernandez OL, et al. Is the Downs and Black scale a better tool to appraise the quality of the studies using virtual rehabilitation for post-stroke upper limb rehabilitation? In: Wright WG, Fluet GG, Subramanian SK, Agmon M, eds. Proceedings of the 2019 International Conference on Virtual Rehabilitation (ICVR); July 22–24, 2019; Tel Aviv, Israel. IEEE Publications. doi: 10.1109/ICVR46560.2019.8994724

49. O’Connor SR, Tully MA, Ryan B, et al. Failure of a numerical quality assessment scale to identify potential risk of bias in a systematic review: a comparison study. BMC Res Notes 2015; 8: 224.

50. Higgins JP, Altman DG, Gotzsche PC, et al. The Cochrane Collaboration’s tool for assessing risk of bias in randomised trials. BMJ 2011; 343: d5928.

51. Sterne JAC HJ, Reeves BC on behalf of the development group for ACROBAT-NRSI. A Cochrane Risk Of Bias Assessment Tool: for Non-Randomized Studies of Interventions (ACROBAT-NRSI), http://www.bristol.ac.uk/population-health-sciences/centres/cresyda/barr/riskofbias/robins-i/acrobat-nrsi/ (2014, accessed October 01, 2023).

52. Hedges LV, Olkin I. Combining estimates of correlation co-efficients. Statistical methods for meta-analysis. Academic Press, Montreal, QC, 2014, pp.224–244.

53. Valentine JC, Pigott TD, Rothstein HR. How many studies do you need? A primer on statistical power for meta-analysis. J Educ Behav Stat 2010; 35: 215–247.

54. Higgins JPT, Thomas J, Chandler J, et al. Cochrane Handbook for Systematic Reviews of Interventions. 2nd ed. Chichester, UK: John Wiley and Sons, 2019.

55. DerSimonian R, Laird N. Meta-analysis in clinical trials. Control Clin Trials 1986; 7: 177–188.

56. Kinney AR, Eakman AM, Graham JE. Novel effect size interpretation guidelines and an evaluation of statistical power in rehabilitation research. Arch Phys Med Rehabil 2020; 101: 2219–2226. DOI: 10.1016/j.apmr.2020.02.017.

57. Chang WH, Bang OY, Shin Y-I, et al. BDNF polymorphism and differential rTMS effects on motor recovery of stroke patients. Brain Stimul 2014; 7: 553–558.

58. Chang WH, Uhm KE, Shin Y-I, et al. Factors influencing the response to high-frequency repetitive transcranial magnetic stimulation in patients with subacute stroke. Restor Neurol Neurosci 2016; 34: 747–755.

59. Kim EJ, Park CH, Chang W, et al. The brain-derived neurotrophic factor Val66Met polymorphism and degeneration of the corticospinal tract after stroke: a diffusion tensor imaging study. Eur J Neurol 2016; 23: 76–84.

60. Park E, Lee J, Chang WH, et al. Differential relationship between microstructural integrity in white matter tracts and motor recovery following stroke based on brain-derived neurotrophic factor genotype. Neural Plast 2020; 2020: 5742421. doi: 10.1155/2020/5742421.

61. Shiner CT, Pierce KD, Thompson-Butel AG, et al. BDNF genotype interacts with motor function to influence rehabilitation responsiveness poststroke. Front Neurol 2016; 7: 69. doi: 10.3389/fneur.2016.00069.

62. Cramer SC, See J, Liu B, et al. Genetic Factors, Brain Atrophy, and Response to Rehabilitation Therapy After Stroke. Neurorehabil Neural Repair 2022; 36: 131–139.

63. Liepert J, Heller A, Behnisch G, et al. Catechol-O-methyltransferase polymorphism influences outcome after ischemic stroke: a prospective double-blind study. Neurorehabil Neural Repair 2013; 27: 491–496.

64. Chang WH, Park E, Lee J, et al. Association between brain-derived neurotrophic factor genotype and upper extremity motor outcome after stroke. Stroke 2017; 48: 1457–1462.

65. Kim BR, Kim HY, Chun YI, et al. Association between genetic variation in the dopamine system and motor recovery after stroke. Restor Neurol Neurosci 2016; 34: 925–934.

66. Subramanian SK, Yamanaka J, Chilingaryan G, et al. Validity of movement pattern kinematics as measures of arm motor impairment poststroke. Stroke 2010; 41: 2303–2308.

67. Kwakkel G, Wagenaar RC, Koelman TW, et al. Effects of intensity of rehabilitation after stroke. A research synthesis. Stroke 1997; 28: 1550–1556.

68. Lang CE, Lohse KR, Birkenmeier RL. Dose and timing in neurorehabilitation: prescribing motor therapy after stroke. Curr Opin Neurol 2015; 28: 549–555.

69. Page SJ, Schmid A, Harris JE. Optimizing terminology for stroke motor rehabilitation: recommendations from the American Congress of Rehabilitation Medicine Stroke Movement Interventions Subcommittee. Arch Phys Med Rehabil 2012; 93: 1395–1399.

70. McHughen SA, Pearson-Fuhrhop K, Ngo VK, et al. Intense training overcomes effects of the Val66Met BDNF polymorphism on short-term plasticity. Exp Brain Res 2011; 213: 415–422.

71. Subramanian S, Chavez M, Gonzalez EA, et al. Estimation of Task Practice Intensity in Individuals with Mild Traumatic Brain Injury [abstract]. Arch Phys Med Rehabil 2021; 102: e52–e53.

72. Cirstea MC, Ptito A, Levin MF. Arm reaching improvements with short-term practice depend on the severity of the motor deficit in stroke. ExpBrain Res 2003; 152: 476–488.

73. Sullivan JE, Crowner BE, Kluding PM, et al. Outcome measures for individuals with stroke: process and recommendations from the American Physical Therapy Association neurology section task force. Phys Ther 2013; 93: 1383–1396. DOI: 10.2522/ptj.20120492.

74. Rehabilitation Measures Database [Internet]. www.sralab.org/, (2023, accessed September 25 2023).

75. Subramanian SK, Cross MK, Hirschhauser CS. Virtual reality interventions to enhance upper limb motor improvement after a stroke: commonly used types of platform and outcomes. Disabil Rehabil Assist Technol 2022;17:107–115.

76. Kwakkel G, Lannin NA, Borschmann K, et al. Standardized measurement of sensorimotor recovery in stroke trials: Consensus-based core recommendations from the Stroke Recovery and Rehabilitation Roundtable. Int J Stroke 2017; 12: 451–461.

77. Weech-Maldonado R. Racial-ethnic differences in nursing home rehabilitation care. In: 6th Annual Medicaid Policy and Research Conference 2007, pp.11–12.

78. Tay MRJ. Hospital readmission in stroke survivors one year versus three years after discharge from inpatient rehabilitation: prevalence and associations in an asian cohort. J Rehabil Med 2021; 53:jrm00208. doi: 10.2340/16501977-2849.

79. Song S, Liang L, Fonarow GC, et al. Comparison of clinical care and in-hospital outcomes of Asian American and white patients with acute ischemic stroke. JAMA Neurol 2019; 76: 430–439.

80. Bhandari VK, Kushel M, Price L, et al. Racial disparities in outcomes of inpatient stroke rehabilitation. Arch Phys Med Rehabil 2005; 86: 2081–2086.

81. Shimizu E, Hashimoto K and Iyo M. Ethnic difference of the BDNF 196G/A (val66met) polymorphism frequencies: the possibility to explain ethnic mental traits. Am J Med Genet B Neuropsychiatr Genet 2004; 126B: 122–123.

82. Domschke K, Deckert J, O’donovan MC, et al. Meta-analysis of COMT val158met in panic disorder: Ethnic heterogeneity and gender specificity. Am J Med Genet B Neuropsychiatr Genet 2007; 144: 667–673.

83. Kumar A, Kumar P, Prasad M, et al. Association between apolipoprotein ε4 gene polymorphism and risk of ischemic stroke: a meta-analysis. Ann Neurosci 2016; 23: 113–121.

84. Zhao F, Yue Y, Jiang H, et al. Shared genetic risk factors for depression and stroke. Prog Neuropsychopharmacol Biol Psychiatry 2019; 93: 55–70.

