## Supplementary material for "Genetic Polymorphisms and Post-Stroke Upper Limb Motor Improvement – A Systematic Review and Meta-Analysis": Supplementary material.pdf

| Study Authors | Chang et al 2014 | Chang et al 2016 | Kim and Cramer 2016 | Kim et al 2016 | shiner et al 2016 | chang et al 2017 | Park et al 2020 | Cramer et al 2022 | Kim et al 2016 COMT | Liepert et al 2013 |
|---|---|---|---|---|---|---|---|---|---|---|
| Criterion # | | | | | | | | | | |
| Hypothesis/aim/objective is clearly described | 1 | 1 | 1 | 1 | 1 | 1 | 1 | 1 | 1 | 1 |
| Main outcomes in introduction or Methods | 1 | 1 | 1 | 1 | 1 | 1 | 1 | 1 | 1 | 1 |
| Patient characteristics clearly described | 1 | 1 | 1 | 1 | 1 | 1 | 1 | 1 | 1 | 1 |
| Interventions of interest clearly described | 1 | 1 | 1 | 1 | 1 | 1 | 1 | 1 | 0 | 1 |
| Principle confounders clearly described | 1 | 1 | 1 | 1 | 0 | 1 | 1 | 1 | 1 | 2 |
| Main findings clearly described | 1 | 1 | 1 | 1 | 1 | 1 | 1 | 1 | 1 | 1 |
| Estimates of random variability provided for main outcomes | 1 | 1 | 1 | 1 | 1 | 1 | 1 | 1 | 1 | 0 |
| All adverse events of intervention reported | 0 | 1 | 0 | 1 | 0 | 0 | 0 | 0 | 0 | 0 |
| Characteristics of patients lost to follow-up described | 0 | 1 | 1 | 1 | 1 | 1 | 1 | 1 | 1 | 1 |
| Probability values reported for main outcomes (if not a specific value e.g. 0.032, then the score is zero) | 1 | 1 | 1 | 1 | 1 | 1 | 1 | 1 | 0 | 1 |

| Study Authors | Chang et al 2014 | Chang et al 2016 | Kim and Cramer 2016 | Kim et al 2016 | shiner et al 2016 | chang et al 2017 | Park et al 2020 | Cramer et al 2022 | Kim et al 2016 COMT | Liepert et al 2013 |
|---|---|---|---|---|---|---|---|---|---|---|
| EXTERNAL VALIDITY | | | | | | | | | | |
| Subjects asked to participate were representative of source population | 0 | 0 | 0 | 0 | 0 | 0 | 0 | 0 | 0 | 0 |
| Subjects prepared to participate were representative of source population | 0 | 0 | 0 | 0 | 0 | 0 | 0 | 0 | 0 | 0 |
| Location and delivery of study treatment was representative of source population | 0 | 0 | 0 | 0 | 0 | 0 | 0 | 0 | 0 | 0 |
| Study participants blinded to treatment | 1 | 0 | 0 | 0 | 0 | 0 | 0 | 1 | 0 | 0 |
| Blinded outcome assessment | 1 | 0 | 0 | 0 | 1 | 0 | 0 | 1 | 1 | 1 |
| Any data dredging clearly described | 1 | 1 | 1 | 1 | 1 | 1 | 1 | 1 | 1 | 1 |
| Analyses adjust for differing lengths of follow-up | 1 | 1 | 1 | 1 | 1 | 1 | 1 | 1 | 1 | 1 |
| Appropriate statistical tests performed | 1 | 1 | 1 | 1 | 1 | 1 | 1 | 1 | 1 | 1 |
| Compliance with interventions was reliable | 1 | 1 | 0 | 1 | 1 | 1 | 1 | 1 | 0 | 1 |
| Outcome measures were reliable and valid | 1 | 1 | 1 | 1 | 1 | 1 | 1 | 1 | 1 | 1 |

| | | | | | | | | | | |
|---|---|---|---|---|---|---|---|---|---|---|
| All participants recruited from the same source population | 1 | 1 | 1 | 1 | 1 | 1 | 1 | 1 | 1 | 1 |
| All participants recruited over the same time period | 1 | 1 | 0 | 0 | 0 | 0 | 0 | 0 | 0 | 0 |
| Study Authors | Chang et al 2014 | Chang et al 2016 | Kim and Cramer 2016 | Kim et al 2016 | shiner et al 2016 | chang et al 2017 | Park et al 2020 | Cramer et al 2022 | Kim et al 2016 COMT | Liepert et al 2013 |
| Participants randomized to treatment(s) | 0 | 0 | 0 | 0 | 1 | 0 | 0 | 1 | 0 | 0 |
| Allocation of treatment concealed from investigators and participants | 1 | 0 | 0 | 0 | 1 | 0 | 0 | 1 | 0 | 1 |
| Adequate adjustment for confounding | 0 | 0 | 1 | 0 | 1 | 1 | 1 | 1 | 1 | 1 |
| Losses to follow-up taken into account | 1 | 1 | 1 | 1 | 1 | 1 | 1 | 1 | 1 | 1 |
| POWER | | | | | | | | | | |
| Sufficient power to detect treatment effect as significance level of 0.05 "Sample sizes have been calculated to detect a significant difference" | 0 | 0 | 0 | 0 | 0 | 0 | 0 | 0 | 0 | 0 |
| TOTAL | **19** | **18** | **16** | **17** | **19** | **17** | **17** | **21** | **15** | **19** |

| | Random sequence generation (selection bias) | Allocation concealment (selection bias) | Blinding of participants and personnel (performance bias) | Blinding of outcome assessment (detection bias) | Incomplete outcome data (attrition bias) | Selective reporting (reporting bias) | Other bias |
|---|---|---|---|---|---|---|---|
| Chang et al 2014 | + | + | + | + | + | + | ? |
| Cramer et al. 2022 | + | + | + | + | + | + | ? |

**Table S1 Risk of bias assessment for non-randomized studies**

| | Description | Liepert et al. 2013 | Chang et al. 2016 | Kim et al. 2016a | Kim et al. 2016b | Kim et al. 2016c | Shiner et al. 2016 | Chang et al. 2017 | Park et al. 2020 |
|---|---|---|---|---|---|---|---|---|---|
| **Bias due to confounding** | | | | | | | | | |
| *1.1 Is confounding of the effect of intervention unlikely in this study? If Y or PY to 1.1: the study can be considered to be at low risk of bias due to confounding and no further signaling questions need be considered* | All potential confounding factors accounted for in the demographics table and/or analysis | Y | Y | Y | Y | Y | Y | Y | Y |
| **Risk of bias judgement** | Since answer to Q1.1 was Y across all studies | **Low** | **Low** | **Low** | **Low** | **Low** | **Low** | **Low** | **Low** |
| **Bias in selection of participants into the study** | | | | | | | | | |
| *2.1. Was selection into the study unrelated to intervention or unrelated to outcome?* | Selection based upon criteria that had nothing to do with outcome or intervention. | Y | Y | Y | Y | Y | Y | Y | Y |
| *2.2. Do start of follow-up and start of intervention coincide for most subjects?* | Information obtained from studies | Y | Y | Y | Y | Y | Y | Y | Y |
| **Risk of bias judgement** | All eligible participants included *and* start of intervention and follow up coincide for all subjects | **Low** | **Low** | **Low** | **Low** | **Low** | **Low** | **Low** | **Low** |

| | | | | | | | | | |
|---|---|---|---|---|---|---|---|---|---|
| **Bias in measurement of interventions** | | | | | | | | | |
| *3.1 Is intervention status well defined?* | Information as provided in the studies. | Y | Y | N | Y | Y | Y | Y | Y |
| *3.2 Was information on intervention status recorded at the time of intervention?* | | Y | Y | NI | Y | Y | Y | Y | Y |
| *3.3 Was information on intervention status unaffected by knowledge of the outcome or risk of the outcome?* | | Y | Y | NI | Y | Y | Y | Y | Y |
| **Risk of bias judgement** | Intervention status is well defined and based solely on information collected at the time of intervention, except where this information was not available | **Low** | **Low** | **NI** | **Low** | **Low** | **Low** | **Low** | **Low** |
| **Bias due to departures from intended interventions** | | | | | | | | | |
| *4.1 Were the critical co-interventions balanced across intervention groups?* | Information as provided in the studies. | Y | Y | NI | Y | Y | Y | Y | Y |
| *4.2. Were numbers of switches to other interventions low?* | | Y | Y | NI | Y | Y | Y | Y | Y |
| *4.3. Was implementation failure minor?* | | Y | Y | NI | Y | Y | Y | Y | Y |
| **Risk of bias judgement** | No bias due to departure from the intended intervention | **Low** | **Low** | **NI** | **Low** | **Low** | **Low** | **Low** | **Low** |

| | | | | | | | | | |
|---|---|---|---|---|---|---|---|---|---|
| | is expected, except where this information was not available | | | | | | | | |
| **Bias due to missing data** | | | | | | | | | |
| *5.1 Are outcome data reasonably complete?* | Information as provided in the studies. | Y | Y | Y | Y | Y | Y | Y | Y |
| *5.2 Was intervention status reasonably complete for those in whom it was sought?* | | Y | Y | NI | Y | Y | Y | Y | Y |
| *5.3 Are data reasonably complete for other variables in the analysis?* | | Y | Y | Y | Y | Y | Y | Y | Y |
| *5.4 Are the proportion of participants and reasons for missing data similar across interventions?* | | Y | Y | NI | Y | Y | Y | Y | Y |
| *5.5 Were appropriate statistical methods used to account for missing data?* | | Y | Y | Y | Y | Y | Y | Y | Y |
| **Risk of bias judgement** | Data were reasonably complete across all studies. | **Low** | **Low** | **Low*** | **Low** | **Low** | **Low** | **Low** | **Low** |
| **Bias in measurement of outcomes** | | | | | | | | | |
| *6.1 Was the outcome measure objective?* | Information as provided in studies. | Y | Y | Y | Y | Y | Y | Y | Y |
| *6.2 Were outcome assessors unaware of the intervention received by study participants?* | Not intervention, but polymorphism status was unknown | Y | Y | NI | Y | Y | Y | Y | Y |

| | | | | | | | | | |
|---|---|---|---|---|---|---|---|---|---|
| *6.3 Were the methods of outcome assessment comparable across intervention groups?* | Information as provided in studies. | Y | Y | Y | Y | Y | Y | Y | Y |
| *6.4 Were any systematic errors in measurement of the outcome unrelated to intervention received?* | | Y | Y | NI | Y | Y | Y | Y | Y |
| **Risk of bias judgement** | The methods of outcome assessment were comparable across groups | **Low** | **Low** | **Low** | **Low** | **Low** | **Low** | **Low** | **Low** |
| **Bias in selection of the reported result** | | | | | | | | | |
| Is the reported effect estimate unlikely to be selected, on the basis of the results, from... | | | | | | | | | |
| *7.1. ... multiple outcome measurements within the outcome domain?* | All studies used different outcomes | Y | Y | Y | Y | Y | Y | Y | Y |
| *7.2 ... multiple analyses of the intervention-outcome relationship?* | Outcomes were assessed at Body structure and/or body function as well as at activity limitation level | Y | Y | Y | Y | Y | Y | Y | Y |
| *7.3 ... different subgroups?* | Only one study had subgroup analyses, information not available for others | NI | NI | NI | NI | NI | Y | NI | NI |
| **Risk of bias judgement** | There is clear evidence that all reported results correspond to all outcomes and analyses. | **Low** | **Low** | **Low** | **Low** | **Low** | **Low** | **Low** | **Low** |
